# Statin Use and Genetically Predicted HMG-CoA Reductase Inhibition in Relation to Clonal Hematopoiesis

**DOI:** 10.64898/2026.07.08.26357595

**Authors:** Paul R Carter, Malgorzata Gozdecka, Sean Wen, Pedro M. Quirós, Samuel M. Lockhart, Monika Dudek, Laura Bond, George Richenberg, Susanna C. Larsson, Daniel Bromage, Jonathan Mitchell, Brian Huntly, Peter Libby, Murray C. H. Clarke, Margarete Fabre, George S. Vassiliou, Stephen Burgess, Siddhartha P. Kar

## Abstract

**Background:** Clonal hematopoiesis (CH) is associated with increased risks of diverse cardiovascular diseases, hematologic malignancies and mortality, yet no preventive therapies are approved. As emerging data implicate lipid pathways in CH pathogenesis, we investigated the association of statin use and genetically proxied inhibition of HMG-CoA reductase (HMGCR) with CH risk, and validated findings using primary peripheral blood mononuclear cells (PBMCs).

**Methods:** We performed an observational analysis of 416,118 UK Biobank participants of European ancestry using multivariable logistic regression to compare CH prevalence among statin users and nonusers. Mendelian randomization (MR) analyses evaluated the causal association of genetically proxied lowering of low-density lipoprotein cholesterol (LDL-C) with risk of CH using two instruments; (i) the lead *HMGCR* variant (rs12916) which proxied LDL-C lowering by statins, and, (ii) 303 genome-wide LDL-C-lowering variants representing polygenic mechanisms. Summary statistics were obtained from the Global Lipid Genetics Consortium genome-wide association study (N = 842,634). Experimentally, primary PBMCs from a *DNMT3A*^R882^ hotspot mutation carrier were cultured in methylcellulose with pravastatin or vehicle control to evaluate colony-forming dynamics.

**Results:** Among 416,118 individuals, 20,488 had CH, including 11,550 with single *DNMT3A*-mutant and 4,375 with single *TET2*-mutant CH. Pre-recruitment statin users had reduced odds of *DNMT3A*-mutant CH (OR=0.93; 95% CI:0.88-0.98; *P*=0.009), driven primarily by associations with *DNMT3A*^R882^–mutant (OR=0.78; 95% CI:0.66-0.92; *P*=0.003), but not *TET2*-mutant CH (OR=1.05; 95% CI:0.97-1.14; *P*=0.20). Similarly, genetically predicted HMG-CoA-reductase inhibition equivalent to a 1 SD reduction in circulating LDL-C levels was associated with lower odds of *DNMT3A*-mutant CH (OR=0.66; 95% CI:0.45-0.95; *P*=0.03) but not *TET2*-mutant CH (OR=1.34; 95% CI:0.76-2.36; *P* = 0.31). By contrast, polygenic estimation of LDL-C lowering was not associated with *DNMT3A*-mutant CH (OR=1.05; 95% CI:0.97-1.14; *P*=0.20), suggesting protective effects were independent of LDL-C lowering per se. Genetically predicted HMG-CoA reductase inhibition had wide effects on blood cell counts and indices, suggesting effects on bone marrow cell dynamics. *In vitro*, pravastatin selectively suppressed colony formation of primary human *DNMT3A*^R882^–mutant relative to wild-type cells (P=0.031).

**Conclusions:** Statin therapy and genetically predicted lifelong inhibition of HMG-CoA reductase were significantly associated with reduced risk of *DNMT3A*-mutant CH, likely via LDL-C-independent mechanisms, which may be specific to *DNMT3A*-mutant CH. This provides a strong rationale for prospective trials evaluating the effect of statins on risk of developing *DNMT3A*-mutant CH, subsequent clonal expansion, and associated clinical sequelae.

**Clinical Perspectives:** *What is new?:*

- This study provides the first evidence that statins may protect against *DNMT3A*-mutant clonal hematopoiesis (CH), with convergent findings of observational analysis of statin users and genetic epidemiology utilizing Mendelian randomization to proxy lifelong HMG-CoA reductase inhibition.
- Observational analyses suggest the protective effect may be strongest against *DNMT3A*^R882^–mutant CH.
- We provide evidence that pravastatin selectively suppresses colony formation of primary human *DNMT3A*^R882^–mutant primary peripheral blood mononuclear cells *in vitro*, suggesting a possible effect on clonal expansion.

*What are the clinical implications?:*

- Although *DNMT3A*-mutant CH is associated with diverse cardiovascular diseases, hematological malignancies and mortality, no preventive treatments are currently established; this study provides a viable avenue for future interventional studies.
- These data establish a strong rationale for prospective trials evaluating whether statins prevent the development or expansion of *DNMT3A*-mutant CH and its cardiovascular consequences.
- Given that statins are widely used with established safety profiles, affordable and have proven cardiovascular benefit, such trials are readily implementable.

## Introduction

Clonal hematopoiesis (CH) is characterized by the age-related expansion of hematopoietic stem cells (HSCs) harboring somatic mutations that confer a fitness advantage. In over 70% of cases these mutations occur in the epigenetic regulators *DNMT3A* or *TET2*. CH is highly prevalent in individuals over 60 years of age, and associates independently with severe clinical sequelae, including hematological malignancies and mortality.^1,2^ The latter is largely driven by an elevated risk for a broad spectrum of cardiovascular diseases, such as heart failure, atrial fibrillation, and accelerated atherosclerosis.^1–8^ Despite these myriad adverse consequences, there are currently no clinical trial-proven pharmacological therapies for the prevention or management of CH.

Statins are used for low-density lipoprotein cholesterol (LDL-C) lowering in cardiovascular disease (CVD) prevention and are among the most commonly prescribed drugs worldwide. Statins act by inhibiting 3-hydroxy-3-methyl-glutaryl-coenzyme A (HMG-CoA) reductase, which is encoded by *HMGCR*. HMG-CoA reductase catalyzes the rate-limiting step in the mevalonate pathway that governs cholesterol biosynthesis. The mevalonate pathway is also responsible for the production of mevalonate-derived isoprenoids, which serve as lipid attachments for the post-translational modification of key intracellular signaling proteins^9^ that play major roles in cell growth and death,^10^ angiogenesis,^11^ and inflammation^12^. Several laboratory- and population-based studies support HMGCR inhibition as a cancer prevention and treatment strategy due to these non-LDL-C-dependent downstream impacts of the mevalonate pathway.^13–15^

Given the cardiovascular benefits of statins, their clinical use has been proposed for the management of cardiovascular risk in CH.^16^ In the English Longitudinal Study of Ageing, statins had greater benefit on cardiovascular primary prevention in CH compared to controls, but also reduced clonal expansion of *TET2*-mutant clones, but these analyses were based on only 400 individuals and 93 *TET2* mutation carriers, respectively.^17^ Beyond CVD, CH drives a greater than 10-fold risk of progression to subsequent myeloid malignancies, with leukemic cells demonstrating high sensitivity to statin-induced apoptosis in preclinical models.^18,19^ This therapeutic potential is supported by encouraging phase I/II trial results combining high-dose pravastatin for acute myeloid leukaemia (AML)^20,21^, alongside large observational datasets demonstrating reduced risk of incident myeloproliferative neoplasms (MPN)^22^, progression to AML^23^, and improved survival in myelodysplastic syndrome (MDS)^23^. Collectively, this motivates an investigation of HMGCR inhibition in the prevention of CH and its sequelae.

Here, we employ a multi-faceted approach to investigate the potential of statins in CH prevention. First, we conduct an observational analysis of over 416,000 UK Biobank (UKB) participants, comparing CH prevalence between statin users and non-users. Second, we leverage a robust *HMGCR* genetic variant to proxy lifelong LDL-C lowering through HMGCR-mediated mechanisms. Using two-sample Mendelian randomization (MR), we evaluated the genetic associations between LDL-C lowering at *HMGCR*, and risk of *DNMT3A*-mutant and *TET2*-mutant CH, as well as on hematological indices. Third, we experimentally evaluate the effect of statins on colony formation by *DNMT3A*-mutant and wild-type mononuclear cells derived from an individual with CH. Together these complementary analyses triangulate a potential benefit of statins in individuals with *DNMT3A*-mutant CH.

## Methods

### Observational analysis of CH risk amongst statin users

Inclusion of UKB participants and CH detection was performed as previously described.^24,25^ Specifically, 416,118 individuals of European descent with whole-exome sequencing (WES) data passing quality control criteria, without pre-recruitment blood cancer diagnosis, and CH data available were included in our study. Somatic variant calling on established driver CH mutations was performed to identify individuals with CH.^26^ Among individuals with CH, only individuals with mutation(s) in one driver gene (mono-gene mutant individuals) were included for downstream association analysis. Self-reported pre-recruitment use of selected medications was ascertained from verbal interviews, namely statin, metformin, sulphonylureas, insulin, thiazolidinediones, meglitinides, and acarbose. Statin users not on any of these diabetic medications were included for downstream association analysis as we have found that metformin reduces risk of *DNMT3A*-mutant CH,^24^ which could confound analyses as statins are often co-prescribed. Specifically, 53,835 statins-only users, 14,307 users on statins and on ≥1 diabetic medication, and 347,976 individuals not on statins were identified.

### Mendelian randomization analyses

We used a two-sample Mendelian randomization design, obtaining genetic associations with LDL-C from one genome-wide association study (GWAS) and with CH from an independent GWAS to evaluate the association between genetically proxied LDL-C and CH risk. Mendelian randomization rests on three main assumptions that relate to the genetic instrument (germline variants used to proxy an exposure): the variants are associated with the exposure (the relevance assumption), the variants are not associated with other risk factors for the outcome (the exchangeability assumption), and the variants affect outcome risk only via the exposure (the exclusion restriction assumption). Two-sample Mendelian randomization is less likely to yield false positive exposure-outcome associations due to weak instrument bias compared to one-sample methods.^27^

### Exposures for MR analyses

This study only used publicly available, deidentified, summary-level GWAS data from previously published studies subjected to review by appropriate ethics committees or institutional review boards and to informed consent procedures in accordance with the Declaration of Helsinki. We proxied HMG-CoA reductase inhibition, using genetic variation at *HMGCR*, the target of statins, associated with lower LDL-C. Specifically, we used the lead variant with the strongest association to LDL-C (rs12916, *P* = 5.6^−404^) within 100kb of *HMGCR* in a GWAS from the Global Lipids Genetics Consortium 2021, with no overlapping UKB participants (GLGC; N = 842,634 individuals of European ancestry).^28^ Moreover, this variant, rs12916, is well established as a genetic proxy for HMGCR inhibition and is known to successfully recapitulate the protective effect of statin use on a range of cardiovascular and metabolic outcomes.^29–33^

Secondary analyses evaluated associations between lower LDL-C proxied by polygenic mechanisms to determine whether associations with CH risk were specific to the *HMGCR* gene region. The polygenic instrument for LDL-C reduction was comprised of 303 independent (*r*^2^ < 0.001) genome-wide variants associated with LDL-C at the genome-wide significance level in the GLGC meta-analysis (without UKB participants).^28^

### Outcomes for MR analyses

Summary-level genetic association data for CH risk were obtained from a previously reported case-control GWAS of CH among European ancestry individuals in the UK Biobank.^25^ Primary analyses focused on *DNMT3A*- and *TET2*-mutant CH risks since these two most commonly mutated CH driver genes together accounted for nearly 80% of individuals with detectable CH (11,836 *DNMT3A*- and 4,951 *TET2*-mutant cases and 386,860 controls without detectable CH). Outcomes for other less common driver genes for which GWAS were available (*ASXL1, PPM1D, TP53, SF3B1, SRSF2, GNB1, IDH2* and *JAK2)* were also studied but given their rarity, analyses were underpowered. As previous GWAS have varied in terms of CH calling methods, associations with *DNMT3A-*mutant CH were replicated in two further UKB-based GWAS of 179,103^2^ (including 5,185 cases) and, 359,088 participants^34^ (including 16,219 cases) Exome sequence data for CH detection were generated using Illumina NovaSeq 6000^35^ and the germline genotypes for the GWAS were obtained from direct array (Applied Biosystems UK BiLEVE Axiom and UK Biobank Axiom arrays) genotyping of blood-derived DNA and imputed with Haplotype Reference Consortium (HRC) and UK10K + 1000 Genomes panel.^36^

Lastly, we explored the associations of genetically predicted HMGCR inhibition with blood cell traits. We obtained summary statistics from the Blood Cell Consortium GWAS meta-analysis of 562,132 individuals of European ancestry.^37,38^ The associations of 15 traits related to red blood cell, white blood cell or platelet indices to genetically predicted *HMGCR* inhibition were studied.

### Statistical Analysis

In observational analyses, logistic regression modelled the association between CH as the outcome and pre-recruitment statin use as the predictor, adjusting for age, sex, smoking, waist-to-hip ratio (WHR)-adjusted body mass index (BMI), and first four genetic principal components.

The primary MR analyses tested for associations between lower LDL-C as genetically predicted by the lead variant at *HMGCR* and risks of *DNMT3A*-mutant and *TET2*-mutant CH. The secondary MR analyses tested for associations between lower LDL-C as genetically predicted across the genome and *DNMT3A*-mutant CH. We also tested for associations between the lead *HMGCR* variant with blood cell traits. Variants were matched across the LDL-C and outcome data sources by rsID and variant-LDL-C and variant-outcome association effect size estimates by effect allele. The Wald ratio method was used for single variant analyses. For secondary polygenic analyses, the association of each variant with CH risk was weighted by its association with LDL-C to generate per variant Wald ratios and the corresponding approximate standard errors estimated by the delta method. The per variant Wald ratio estimates were combined using random-effects inverse-variance weighted meta-analysis.^39^

For the *HMGCR* instrument, the exclusion restriction assumption was assessed by testing for associations with smoking initiation (defined as ever having smoked regularly) and leukocyte telomere length (LTL), both of which are strong CH risk factors.^2^ Summary-level genetic association data for smoking initiation and LTL were obtained from previously published large-scale GWAS.^40,41^ Lack of associations were considered evidence that the LDL-C lowering variants were unlikely to influence CH risk through these traits.

This study is reported in accordance with the “strengthening the reporting of observational studies in epidemiology using Mendelian randomization” (STROBE-MR) guidelines.^42^ All statistical tests were two-sided and *P* < 0.05 was considered statistically significant. The odds ratios (OR) for CH risk from MR analyses were scaled to reflect the equivalent of 1 SD reduction in circulating LDL-C levels. All MR analyses were conducted in R (version 4.2.2) using the TwoSampleMR (version 0.5.6) package.^43^

### Validation of effect of statins on primary human cells

#### Colony plating assay

Human primary *DNMT3A*^R882^ peripheral blood mononuclear cells were plated at ∼300,000/well in 6-well plates, in methylcellulose semi-solid medium (Stem Cell Technologies, H4034) with 400μM pravastatin (Sigma, P4498) or vehicle control. Colonies were counted and picked at day 13 post plating.

#### Colony genotyping

Colonies were collected into DirectPCR Lysis Reagent (Viagen, 401-E) and processed according to manufacturer instructions. Lysed DNA was used for *DNMT3A* genotyping with the following primers; F: 5’-CTGAGTGCCGGGTTGTTTAT-3’, R: 5’-GGAAGGGAGCTTGGTTTTGT-3’. PCR was performed with REDTaq ReadyMix PCR Reaction Mix (Sigma, R2523) using the following conditions: initial denaturation at 95 °C for 1 min, 35 cycles at 95°C for 15 s, annealing at 57°C for 15 s and elongation at 72°C for 15 s. Final elongation was performed at 72°C for 10 min. The presence of *DNMT3A*^R882^ generates additional restriction sites for AluI enzymes, thus PCR products were digested with the AluI restriction enzyme (R0137S) for 1h at 37°C and visualized on 2.5% agarose gel.

## Results

### Statin users in UK Biobank have reduced risk of DNMT3A CH, driven by associations with DNMT3A^R882^ CH

Statins have been proposed to reduce *TET2* clonal expansion^17^ and may prevent myeloid malignancies for which CH is a precursor.^20–23,44^ We therefore assessed whether statin use at the time of UK Biobank recruitment was associated with concurrent CH status (*n* = 53,835 statin users and 347,976 controls), excluding individuals on diabetic medications (Figure 1; Supplemental Tables 1-2). Statin users had reduced risk of *DNMT3A*-mutant CH (OR = 0.93; 95% CI: 0.88-0.98; *P* = 0.009; *n* = 11,550 cases) which was driven by a specific and more pronounced association with *DNMT3A^R^*^882^ CH (OR = 0.78; 95% CI: 0.66-0.92; *n* = 1,454 cases; *P* = 0.003), with the protective association not seen for non-R882 *DNMT3A* CH (OR = 0.95; 95% CI: 0.90-1.01 *n* = 10,096 cases; *P* = 0.089). Statin users also had lower risk of *SF3B1*-mutant CH (OR = 0.65; 95% CI: 0.43-1.00; n = 172 cases; *P* = 0.048) but no associations were found with CH overall (OR = 0.97; 95% CI: 0.93-1.01; n = 20,488 cases; *P* = 0.144) or with other gene drivers, including *TET2*-mutant CH (OR = 1.04; 95% CI: 0.96-1.13; *n* = 4,375 cases; P = 0.325).

**Figure 1.**
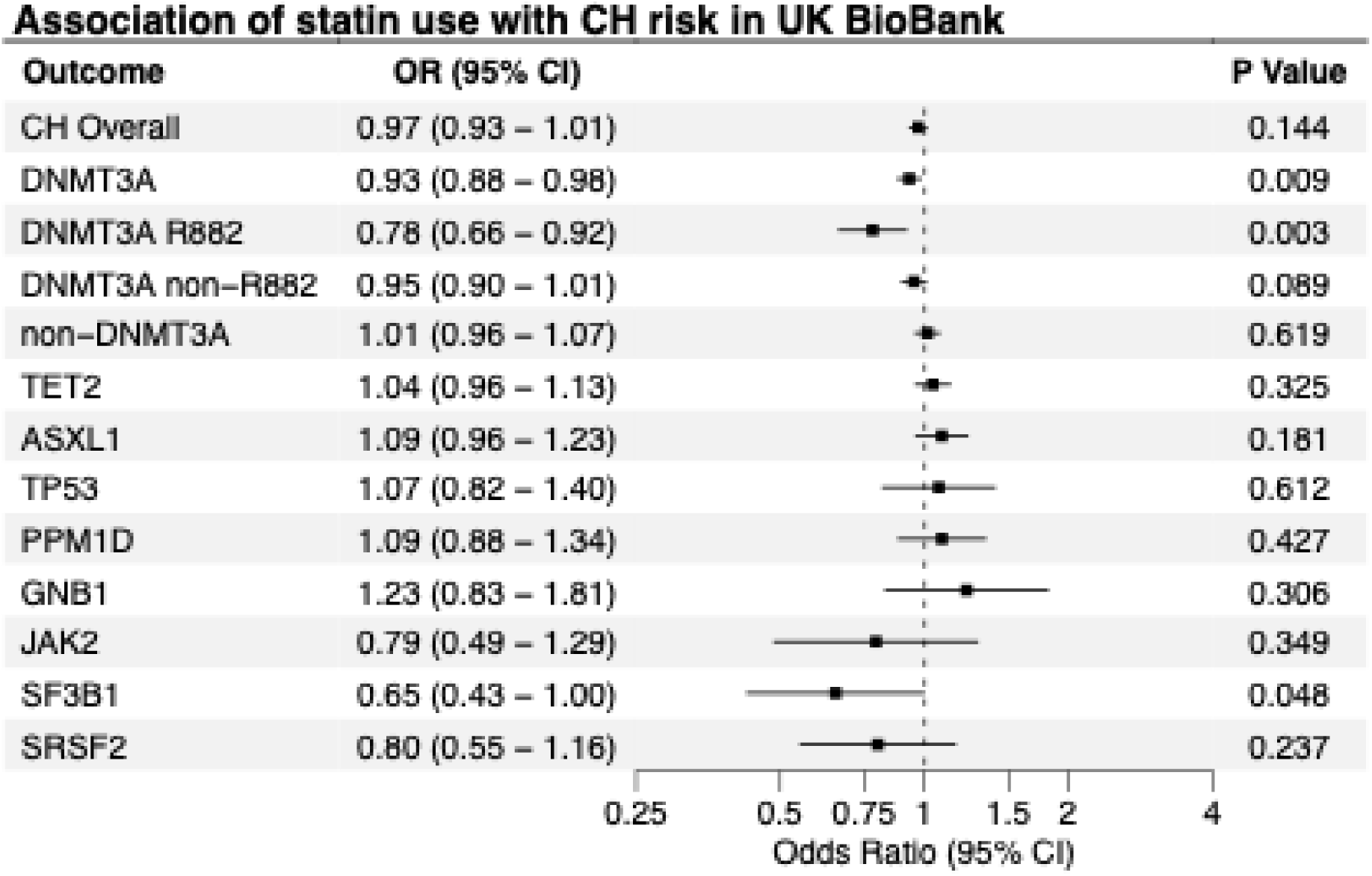
Observational analysis of CH risk amongst pre-recruitment statin users in UK Biobank. OR indicates odds ratio; CI, confidence interval; and CH, clonal hematopoiesis. Data markers indicate the point estimate for the OR in logistic regression analyses accounting for age, sex, smoking, waist-to-hip ratio-adjusted body mass index, and first four genetic principal components. Error bars indicate the 95% confidence interval. Only gene-specific driver CH with ≥10 statin users were included here.

### Genetically predicted HMG-CoA reductase inhibition is protective for DNMT3A CH, but not TET2 CH

*DNMT3A* and *TET2* are the two most commonly mutated CH driver genes, with opposing epigenetic roles and have distinct germline susceptibility architectures (most notably the *TCL1A* locus) and clonal trajectories.^2,45,46^ To further investigate the protective association between HMG-CoA reductase inhibition and CH, and to mitigate biases of observational analyses, we performed genetic analysis. CH outcomes were from GWAS of 406,826 European individuals in UKB (Figure 2; Supplemental Table 2).^25^ Genetically predicted LDL-C-lowering equivalent to a 1 SD reduction in LDL-C per T allele of the *HMGCR* SNP rs12916 was significantly associated with a reduced risk of *DNMT3A*-mutant CH (OR = 0.66; 95% CI: 0.45-0.95; *P* = 0.03; Figure 2; 11,836 cases and 386,860 controls). However, there was no association with *TET2*-mutant CH (OR = 1.34; 95% CI 0.76-2.36 per SD lower LDL-C; *P* = 0.31; Figure 2; 4,951 cases and 386,860 controls). Genetically predicted LDL-C lowering per T allele of the *HMGCR* SNP rs12916 was not associated with risk of CH overall (OR = 0.88; 95% CI 0.66-1.18 per SD lower LDL-C; *P* = 0.40; 19, 966 cases and 386,860 controls), or of any other subtypes (Supplemental Figure 1), including *SF3B1*-mutant CH (OR = 8.84; 95% CI 0.57-137.7 per SD lower LDL-C; *P* = 0.12; 207 cases and 386,860 controls), but analyses of other drivers were underpowered due to their rarity.

**Figure 2.**
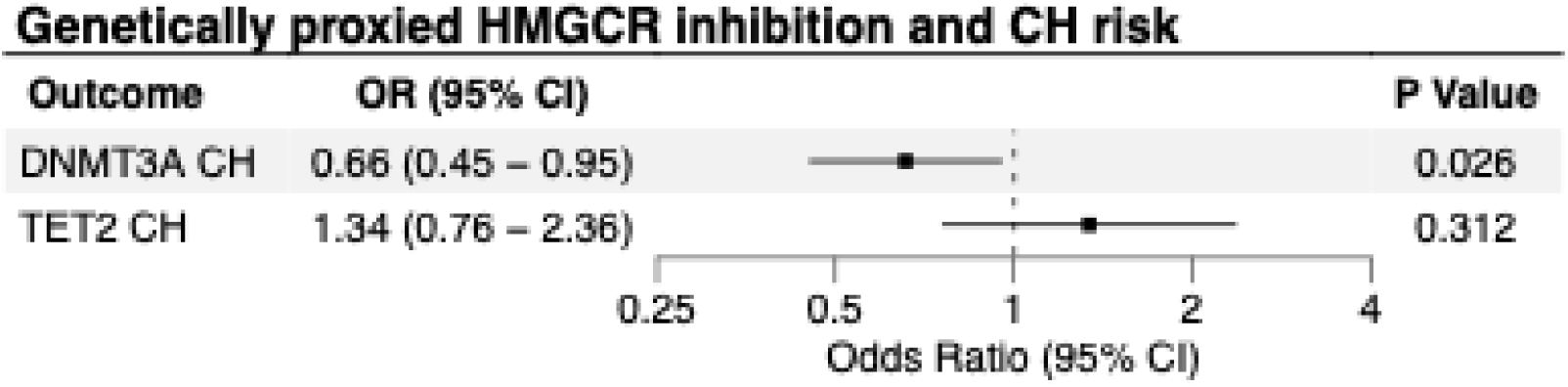
Mendelian randomization estimates of the association between circulating LDL-C level predicted by variation at *HMGCR (rs12916)* and *DNMT3A*-mutant and *TET2*-mutant CH risk. OR indicates odds ratio; CI, confidence interval; LDL-C, low-density lipoprotein cholesterol; and CH, clonal hematopoiesis. Data markers indicate the point estimate for the OR per 1 SD reduction in LDL-C based on the Wald ratio method. Error bars indicate the 95% confidence interval.

Previous CH GWAS have differed in their definitions of CH and in variant calling and filtering pipelines, potentially leading to heterogeneity in genetic associations. To assess the robustness of our findings, we repeated the analyses in two independent UKB-based CH GWAS (Supplemental Figure 2; Supplemental Table 2).^2,34^ Consistent with our primary results, genetically predicted LDL-C lowering equivalent to a 1 SD reduction in LDL-C per T allele of the HMGCR variant rs12916 was associated with a reduced risk of *DNMT3A*-mutant CH in both the 179,103 participant GWAS^2^ (OR = 0.41; 95% CI 0.23-0.71 per SD lower LDL-C; *P* = 0.001) and the 359,088 participant GWAS^34^ (OR = 0.65 ; 95% CI 0.47-0.89 per SD lower LDL-C; *P* = 0.007).

### Genetic inhibition of HMGCR profoundly impacts blood cell counts

We hypothesized that if statins influence HSC homeostasis, genetically predicted HMG-CoA reductase inhibition would have wider effects on blood cell counts and indices. To test this hypothesis, we conducted MR analyses using GWAS meta-analysis data for 15 hematological traits from up to 562,132 individuals from the UKB and the Blood Cell Consortium (Figure 3; Supplemental Table 2)).^37,38^ Indeed, HMG-CoA reductase inhibition as genetically predicted by the T allele of rs12916 (equivalent to a 1 SD reduction in circulating LDL-C level) was significantly associated with reduced platelet count (OR = 0.80; 95% CI: 0.76-0.84; *P* = 2.6 × 10^−17^), hemoglobin level (OR = 0.94; 95% CI: 0.90-0.99 ; *P* = 0.02) and mean corpuscular hemoglobin concentration (OR = 0.90; 95% CI: 0.86-0.96; *P* = 4.2 × 10^−4^); as well as increased red cell distribution width (OR = 1.11; 95% CI: 1.06-1.17; *P* = 5.8 × 10^−5^), monocyte count (OR = 1.10; 95% CI: 1.05-1.16; *P* = 2.0 ×10^−4^) and mean platelet volume (OR = 1.17; 95% CI: 1.11-1.23; *P* = 1.5 × 10^−8^).

**Figure 3.**
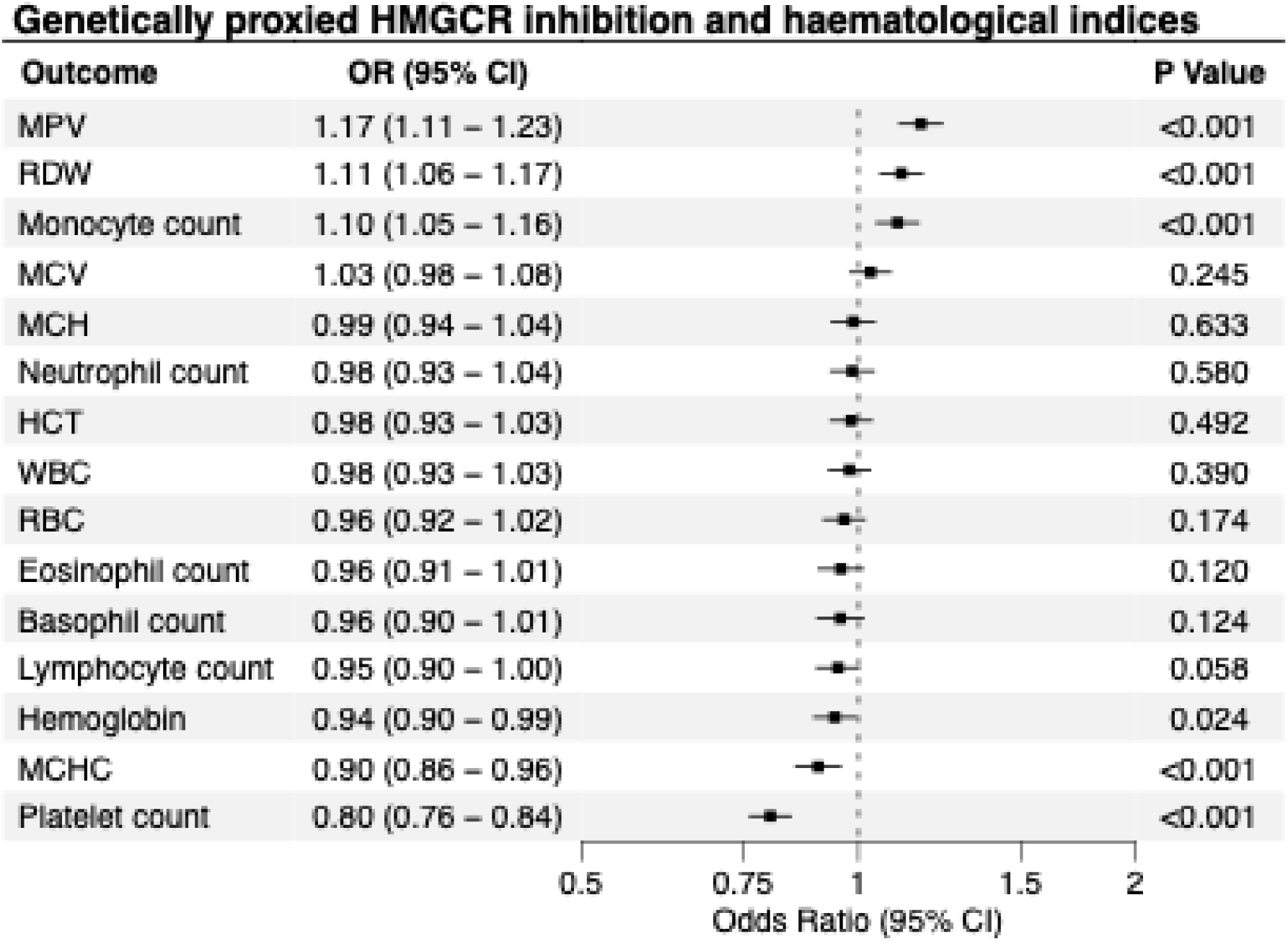
Mendelian randomization estimates of the association between circulating LDL-C level predicted by variation at *HMGR (rs12916) and blood cell traits*. OR indicates odds ratio; CI, confidence interval; LDL-C, low-density lipoprotein cholesterol; WBC, white blood count; Neut, neutrophil count; Mono, monocyte count; Lymph, lymphocyte count; Eos, eosinophil count; Baso, basophil count; Hb, hemoglobin; HCT, hematocrit; RBC, red blood cell count; MCV, mean corpuscular volume; MCH, mean corpuscular hemoglobin; MCHC, mean corpuscular hemoglobin concentration; RDW, red cell distribution width; PLT, platelet count and MPV, mean platelet volume. Data markers indicate the point estimate for the OR per 1 SD reduction in LDL-C based on the Wald ratio method. Error bars indicate the 95% confidence interval.

### *Genetic associations of HMGCR* inhibition with *DNMT3A* CH are unlikely to be mediated by LDL-C, smoking initiation or leukocyte telomere length

Next, we investigated whether protective associations on *DNMT3A*-mutant CH were specific to genetic HMGCR inhibition rather than a generalized benefit of lowering circulating LDL-C. Lower LDL-C level, predicted by a polygenic instrument comprised of 303 independent (*r*^2^ < 0.001) variants^28^ across the genome each associated with blood LDL-C at genome-wide significance, was not associated with *DNMT3A*-mutant CH (OR = 1.05; 95% CI: 0.97-1.14 per SD lower LDL-C; *P* = 0.20; Supplemental Table 3). This suggested that genetically predicted LDL-C lowering more generally was not the driver of the protective effect on *DNMT3A*-mutant CH and that this protection was likely due to mechanisms specific to HMG-CoA reductase inhibition.

Finally, we explored whether genetically predicted HMG-CoA reductase inhibition was associated with smoking initiation (ever smoked versus never smoked)^40^ and LTL^41^, which are both established risk factors for *DNMT3A*-mutant CH (Figure 4; Supplemental Table 2).^2^ Genetic inhibition of *HMGCR* predicted by rs12916 was not significantly associated with smoking initiation (OR = 1.08; 95% CI: 0.97-1.20; *P* = 0.14) or LTL (OR = 1.01; 95% CI: 0.95-1.07; *P* = 0.76) suggesting these are not genetic confounders in the association with *DNMT3A-mutant* CH.

**Figure 4.**
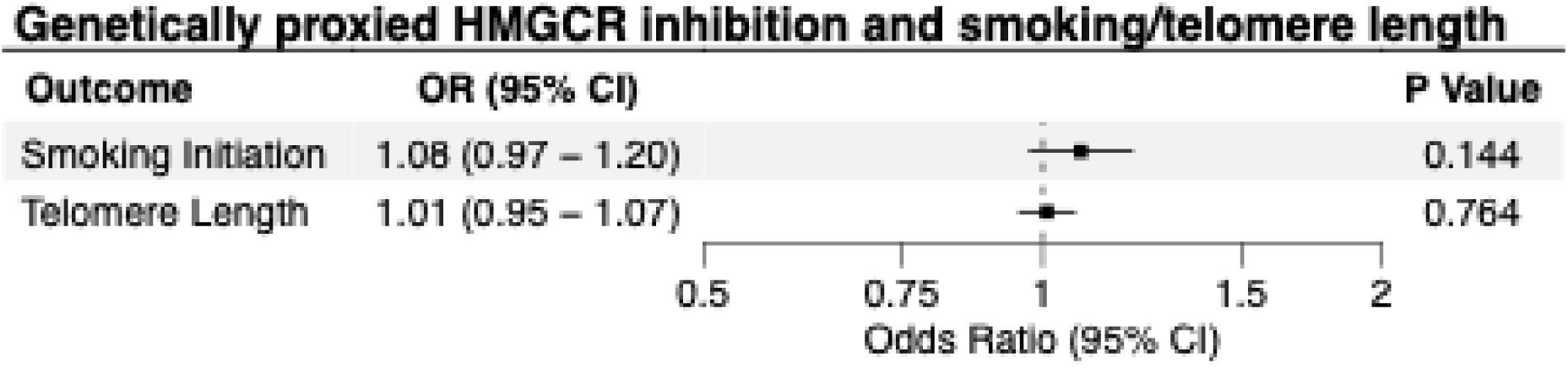
Mendelian randomization estimates of the association between circulating LDL-C level predicted by variation at *HMGCR (rs12916)* and smoking initiation and leukocyte telomere length. OR indicates odds ratio; CI, confidence interval and LDL-C, low-density lipoprotein cholesterol. Data markers indicate the point estimate for the OR per 1 SD reduction in LDL-C based on the Wald ratio method. Error bars indicate the 95% confidence interval.

### Pravastatin suppresses the expansion of primary human DNMT3A-mutant clonal colonies in vitro

As our observational analyses suggested that statins may exert a specific protective effect against *DNMT3A*^R882^ hotspot mutations, we conducted experimental studies with primary human cells to triangulate our MR findings and assess the impact of statins on colony formation in cells harboring this mutation. We plated human blood mononuclear cells from a *DNMT3A*^R882^ mutation carrier (variant allele fraction = 12%) for colony formation in semisolid media in the presence of pravastatin or vehicle control for 13 days (Figure 5a). Sanger sequencing of pooled plate DNA showed a decrease in the proportion of *DNMT3A*^R882^ versus wild type DNA upon pravastatin treatment (Figure 5b-c). Furthermore, genotyping of 96 individual colonies per condition confirmed statistically significant decrease in the proportion of *DNMT3A*^R882^ versus wild type colonies with pravastatin treatment (Figure 5d).

**Figure 5.**
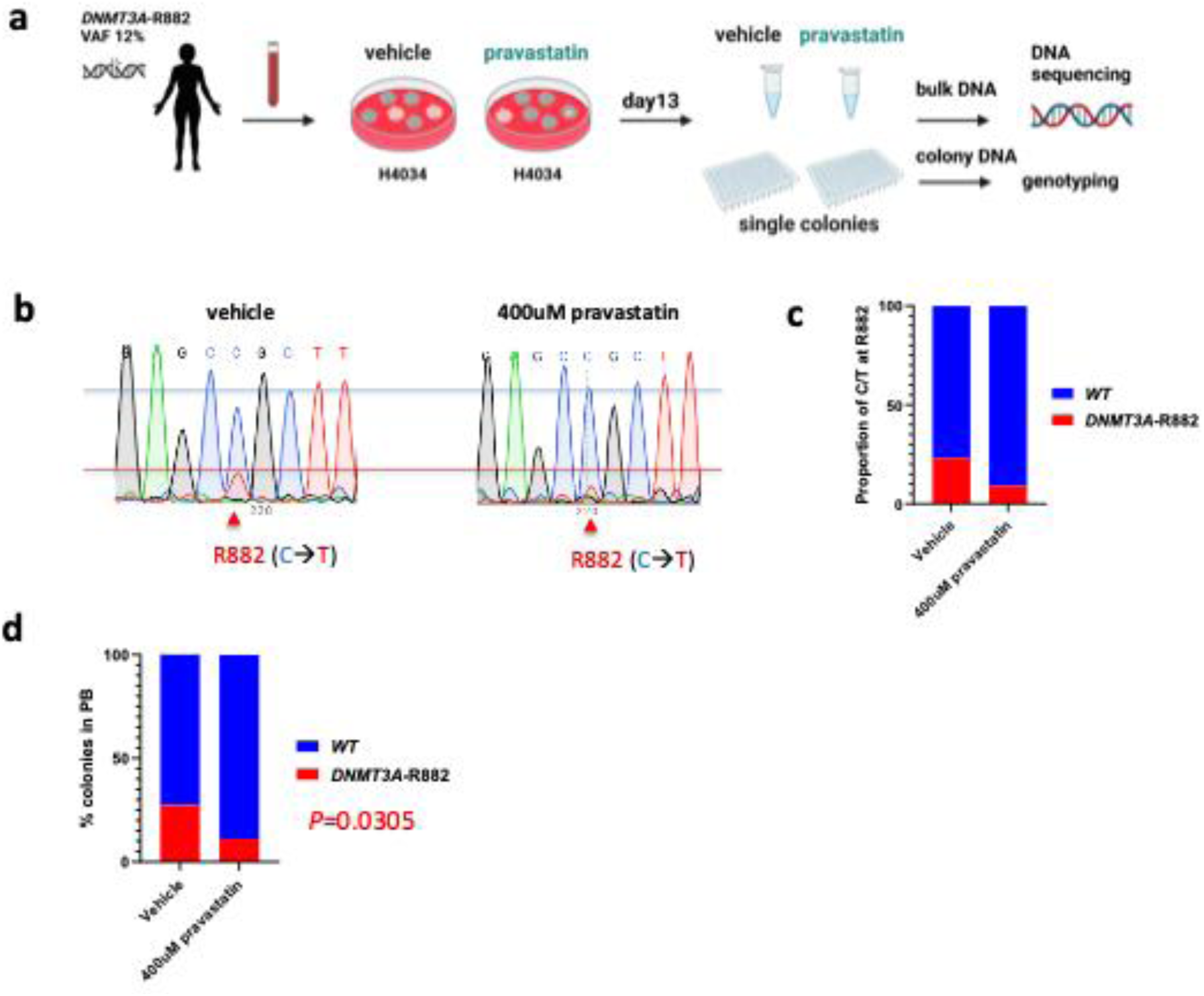
Pravastatin reduces colony formation by *DNMT3A*^R882^–mutant cells from a patient with *DNMT3A* CH. (a) Experimental strategy for testing the effect of pravastatin on *DNMT3A*^R882^ cell clonal growth (vs WT). Schema was created using BioRender. (b-d) Sanger sequencing of DNA collected from bulk colonies (b) and quantification of C/T proportion from Sanger sequencing data (c) Comparison of the proportion of WT versus *DNMT3A*^R882^ colonies in pravastatin-versus vehicle-treated cells (d). *P* by two-sided Chi-square test performed on colony numbers.

## Discussion

This multi-modal study provides convergent evidence that statins specifically protect against *DNMT3A*-mutant CH, with consistent signals from observational data in statin users, MR of genetically proxied statin effects, and *in vitro* colony formation assays. We provide the first evidence of a protective association between LDL-C lowering germline variation in the *HMGCR* region and the risk of developing *DNMT3A*-mutant CH risk - the most common CH driver. Proxying LDL-C reduction via rs12916, the *HMGCR* variant with the strongest association to LDL-C, predicted a 34% decrease in risk of *DNMT3A*-mutant CH per 1 SD reduction. There was little evidence that the association was influenced by genetic confounding from smoking or longer LTL, which are both known risk factors for CH. As cholesterol lowering *HMGCR* variants are proxies for statin therapy, this implies that these widely prescribed and safe medications have the potential to prevent *DNMT3A*-mutant CH. In line with this, statin use amongst UKB participants was associated with a reduced risk of *DNMT3A*-mutant CH, which was specific to the *DNMT3A*^R882^ hotspot mutation which confers markedly elevated risks of leukemic progression. Consistent with this, pravastatin preferentially suppressed formation of *DNMT3A*^R882^ colonies, compared to wild-type colonies formed from human blood mononuclear cells suggesting an effect on clonal expansion. We also find that genetically mimicked action of statins was associated with widespread alterations in blood indices, suggesting potential effects on bone marrow cell dynamics.

Hematopoietic somatic mutations in the tumor suppressor *DNMT3A* have been suggested to drive diverse cardiovascular sequalae including heart failure^6,7^, atherosclerotic disease^3,8^ and atrial fibrillation^2,4,5^, which is thought to be due to augmented pro-inflammatory actions of mutant myeloid cells. They also have a seminal role in CH and the full spectrum of myeloid malignancies, occurring in >50% individuals harboring CH, ∼10% of MDS and MPN cases, and ∼25% of AML cases.^47^ *DNMT3A*-mutant CH can be detected in the blood years before the diagnosis of AML and frequently represents the “first hit” enroute to *NPM1* or *FTL3* mutated AMLs, with the latter representing the downstream driver mutations required for final neoplastic transformation.^48,49^ Notably, *DNMT3A*^R882^ CH confers markedly greater risk of progression to AML^50^. In experiments using cells from a *DNMT3A*^R882^ carrier, statins preferentially suppressed the formation of *DNMT3A*^R882^ colonies relative to wild-type, suggesting they may limit the clonal fitness and expansion of *DNMT3A*-mutant cells. While this does not conclusively demonstrate that HMGCR inhibition has an impact on clonal fitness after a *DNMT3A* mutation has already been acquired, given the increasing interest in using CH, and clone size, to predict future risk of cardiovascular events or hematological cancer, it raises the intriguing possibility that statins may enable early preventive intervention in these settings.

We further found no association between lower LDL-C proxied by variants genome-wide and *DNMT3A*-mutant CH. This result suggests that the protective effect on CH risk derives from mechanisms that correlate with *HMGCR*-specific LDL-C lowering activity independent of LDL-C lowering itself. Downstream of the common step of conversion of HMG-CoA to mevalonate that is catalyzed by HMG-CoA reductase, the mevalonate pathway diverges into distinct arms leading to cholesterol synthesis on one hand and protein geranylation/farnesylation (forms of post-translational modification) on the other. Indeed, Xia et al^51^ have previously shown that lovastatin induced apoptosis in human AML cells, with the cytotoxic effect of lovastatin was reversed by the addition of geranlygeranylpyrophosphate but not by squalene or cholesterol, and the effects of lovastatin were mimicked by geranylgeranyltransferase inhibition. More recently Krosl et al^19^ have shown that inhibition of vesicular trafficking governed by RAB protein geranylgeranylation may be the key vulnerability mediating the anti-AML effect of atorvastatin. Thus, pre-clinical research in AML and other cancers^13^ is in line with our suggestion that genetically proxied HMGCR inhibition decreases CH risk by mechanisms not directly dependent on LDL-C. Our MR results here also parallel other MR findings in oncology wherein genetically predicted HMGCR inhibition but not lower LDL-C more generally associated with solid tumor risk.^15^ Moreover, statins are known to have multiple anti-inflammatory effects^52^ and inflammation can contribute to forms of CH,^53^ and this relationship may, at least in part, also explain this result.

The MR estimates of HMG-CoA reductase inhibition and risk of CH identified here represent the predicted effect of lifelong reduction in LDL-C via HMGCR inhibition. Qualitatively, MR studies involving *HMGCR* variants have shown a remarkable ability to recapitulate the efficacy^54^ and adverse event^55^ profiles of statins in clinical trials. Quantitatively, the shorter term pharmacological HMGCR inhibition using statins in medium-term trials is only able to achieve a third of the reduction in coronary artery disease risk that is predicted by lifelong genetic HMGCR inhibition per unit lowering of LDL-C.^56^ Interestingly, our observational analyses in UKB, which better estimate shorter-term clinical use, revealed a 7% reduction in *DNMT3A*-mutant CH amongst statin users - suggesting a similar ratio between short-term and life-long statin use may exist in CH prevention as it does for coronary artery disease. The combination of our genetic and observational findings provides a strong rationale supporting a trial of statins to decrease CH risk, and potentially even CH progression and sequelae.

### Limitations of this study

Our results are based on individuals of European ancestry in the UKB to reduce the risk of population stratification affecting the MR estimates. Although the association between inherited genetic variation at *HMGCR* and LDL-C is also observed in other ancestries^28,57^, it remains to be seen whether our findings can be generalized to other ancestral groups globally. Furthermore, MR analyses for non-*DNMT3A*/*TET2* CH drivers were underpowered, and as such the associations of genetically predicted HMGCR inhibition with these subtypes need to be studied in larger GWAS when available, particularly as we demonstrate a possible protective effect of statin use on *SF3B1*-mutant CH in observational analyses.

In conclusion, as CH is now widely regarded as a risk factor for numerous diseases, blood and solid cancers and overall mortality, there is a compelling need to identify interventions to prevent its development and progression. Here we show that statin use and genetically predicted HMGCR inhibition are both associated with decreased risk of *DNMT3A*-mutant CH. Further research, including clinical trials, will be required to establish whether HMGCR inhibition by statins indeed prevents the acquisition and expansion of CH mutations and their disease consequences.

## Acknowledgements

The authors thank investigators involved in producing the publicly available data from the UKB, and the BCX and GLGC consortia used to perform this work. Data from the UKB resource were accessed under approved application numbers 68601, 65851, and 26041 for this study.

## Author Contributions

Conceptualization: PRC, SK; Methodology: PRC, SK, SB, MG, SW, GV; Software: PRC, SK, SW MG; Formal Analysis: PRC, SK, SW, MG ; Investigation: PRC, MG, SW, DB, MD, LB, SL, ; Data Curation: PRC, MG, SW, PQ, SK, MF; Writing - Original Draft Preparation: PRC, SK, MG, SW; Writing - Review & Editing: All authors; Visualization: PRC, MG, SW; Supervision: SK, SB, GV, JM, BH, MF, PL, MCHC; Funding Acquisition: PRC, SK, SB, GV, BH, MCHC.

PRC had full access to all of the data in the study and takes responsibility for the integrity of the data and the accuracy of the data analysis.

## Data Availability Statement

The UKB CH data sets used in primary analyses are publicly available at the GWAS Catalog (https://www.ebi.ac.uk/gwas/downloads/summary-statistics) with accession numbers GCST90435354 (*DNMT3A*-mutant CH) and GCST90435355 (*TET2*-mutant CH). UKB-based *DNMT3A*-mutant CH GWAS used in replication analyses are also available from GWAS Catalog with accession numbers GCST90102619 (Kar et al) and GCST90165271 (Kessler et al). The GLGC data sets used are publicly available at https://csg.sph.umich.edu/willer/public/glgc-lipids2021. The Blood Cell Consortium (BCX) data sets used are publicly available from the Lettre Lab at: http://www.mhi-humangenetics.org/en/resources. The smoking initiation (ieu-b-4877) and telomere length (ieu-b-4879) summary statistics are accessible via the MRC IEU OpenGWAS at https://gwas.mrcieu.ac.uk.

## Abbreviations

AML: Acute myeloid leukemia
BCX: Blood Cell Consortium
BMI: Body mass index
CH: Clonal hematopoiesis
CVD: Cardiovascular disease
DNMT3A: DNA methyltransferase 3 alpha
FTL3: Fms-like tyrosine kinase 3 gene
GLGC: Global Lipid Genetics Consortium
GWAS: Genome-wide association study
HMG-CoA: 3-hydroxy-3-methyl-glutaryl-coenzyme A
HMGCR: 3-hydroxy-3-methyl-glutaryl-coenzyme A reductase gene
HSCs: Hematopoietic stem cells
IVW: Inverse-variance weighted
LDL-C: Low-density lipoprotein cholesterol
LTL: Leukocyte telomere length
MCH: Mean corpuscular hemoglobin
MCHC: Mean corpuscular hemoglobin concentration
MDS: Myelodysplastic syndromes
MPN: Myeloproliferative neoplasm
MR: Mendelian randomization
NPM1: Nucleophosmin 1 gene
OR: Odds ratio
PCR: Polymerase chain reaction
PLT: Platelet count
RDW: Red cell distribution width
SF3B1: Splicing factor 3b subunit 1 gene
SNP: Single nucleotide polymorphism
TET2: Ten-eleven translocation 2
UKB: United Kingdom Biobank
WES: Whole-exome sequencing
WHR: Waist-to-hip ratio
WT: Wild-type

---

**Supplemental Figure 1.**
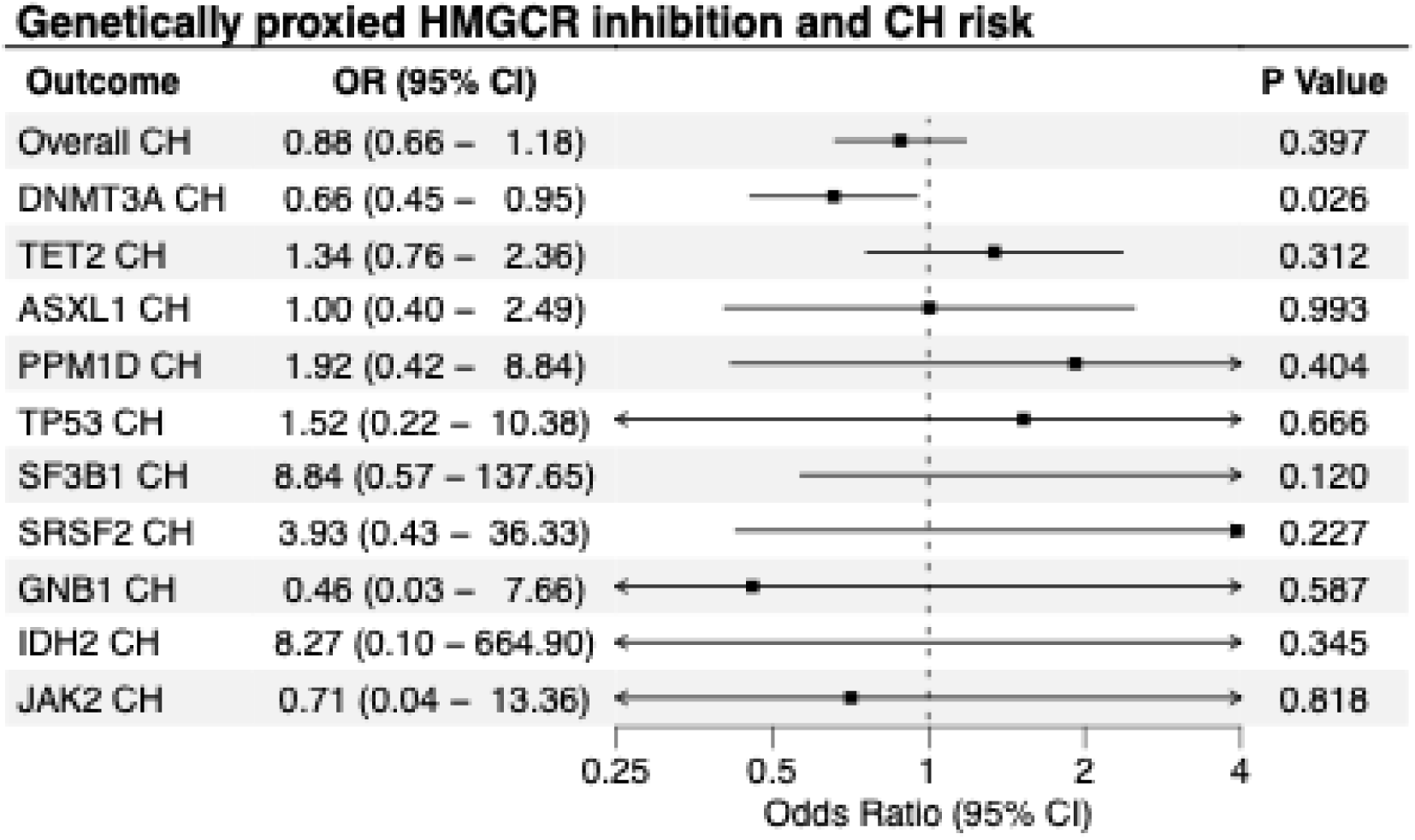
Mendelian randomization estimates of the association between circulating LDL-C level predicted by variation at *HMGCR (rs12916)* and *risk of* CH overall and subtypes. OR indicates odds ratio; CI, confidence interval; LDL-C, low-density lipoprotein cholesterol; and CH, clonal hematopoiesis. Data markers indicate the point estimate for the OR per 1 SD reduction in LDL-C based on the Wald ratio method. Error bars indicate the 95% confidence interval.

**Supplemental Figure 2.**
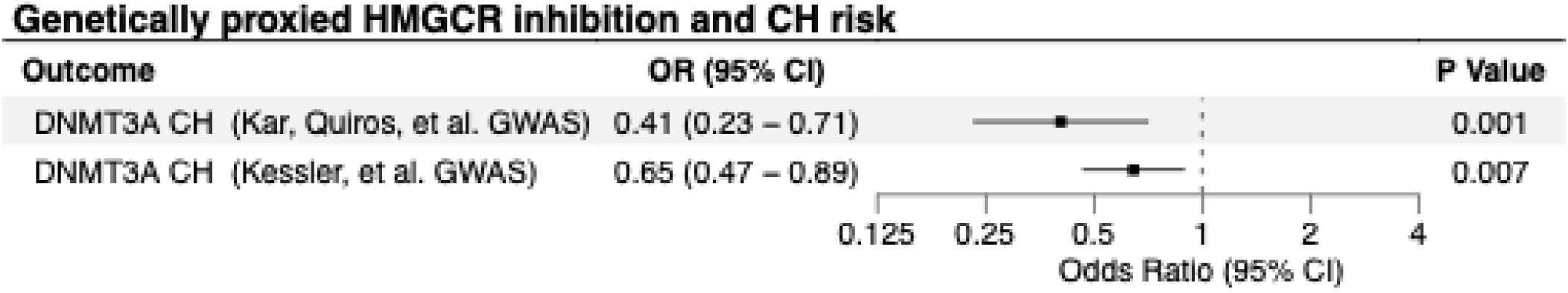
Mendelian randomization estimates of the association between circulating LDL-C level predicted by variation at *HMGCR (rs12916)* and *DNMT3A*-mutant CH in replication analyses using alternative outcome GWAS. OR indicates odds ratio; CI, confidence interval; LDL-C, low-density lipoprotein cholesterol; and CH, clonal hematopoiesis. Data markers indicate the point estimate for the OR per 1 SD reduction in LDL-C based on the Wald ratio method. Error bars indicate the 95% confidence interval.

